# Leveraging Machine Learning for Developing and Validating a Neonatal Acute Kidney Injury Prediction Model (NEPHRO): A Comprehensive Evidence-Based Neonatal AKI Risk Stratification Tool

**DOI:** 10.1101/2025.06.12.25329508

**Authors:** Tahagod Mohamed, Sven Bambach, John David Spencer, Laura Rust, Shama Patel, Jacqueline Magers, Javier A. Neyra, F. Perry Wilson, Xia Ning, Jason Newland, Steve Rust, Jonathan L. Slaughter

## Abstract

**Background:** Acute kidney injury (**AKI**) is a serious and common complication among critically ill neonates. Preventing or treating AKI early requires timely prediction, but current tools to forecast AKI in neonates are limited. We proposed that machine learning could help predict AKI by analyzing routinely collected clinical data.

**Methods:** We conducted a retrospective analysis of 8,059 critically ill neonates admitted to our level IV neonatal intensive care unit (**NICU**) from January 2017 to December 2021 for development and cross-validation (n=5,443, 68%), and from January 2022 to June 2024 (n=2,616, 32%) for temporally validating a neonatal AKI predictive model. Risk factors for model input were identified from the literature, and data were extracted from electronic health records. According to the neonatal modification of kidney disease: Improving Global Outcomes criteria, an increase in serum creatinine (**SCr**) and/or a decrease in urine output (**UOP**) defines neonatal AKI. We trained a machine learning model using a least absolute shrinkage and selection operator (**LASSO**) algorithm to develop and validate a predictive AKI model. The area under the receiver operating characteristic curve (**AUROC**) and F1-scores evaluated the model’s performance.

**Findings:** Among 206,220 NICU patient days, AKI occurred in 881 (11%) neonates. Using 27 potential AKI risk factors, LASSO identified key AKI predictors: fluid balance, hypotension requiring vasopressors, invasive ventilation, sepsis, surgical procedures, and congenital kidney and urinary tract anomalies. The model predicted the occurrence of a critical SCr increase or UOP decrease over the next 48 hours, with an AUROC of 0.814 (95% CI: 0.787-0.843) in development and 0.815 (0.795-0.834) in validation datasets.

**Interpretation:** We developed a machine learning-based model that reliably predicted neonatal AKI before it became clinically apparent by conventional parameters. By identifying high-risk neonates earlier, timely interventions can be deployed to improve outcomes.

## INTRODUCTION

Acute kidney injury (**AKI**) is an abrupt reduction in kidney function that can impact up to 50% of critically ill neonates, particularly preterm infants [1, 2]. AKI is diagnosed by either an increase in serum creatinine (**SCr**) and/or a decrease in urine output (**UOP**) as per the neonatal modification of Kidney Disease Improving Global Outcomes (**nm-KDIGO**) criteria [1]. AKI causes electrolyte derangement, acid-base disturbances, fluid overload, respiratory failure, more extended hospital stays, and an eight-fold increase in mortality during hospitalization in the neonatal ICU (**NICU**) [1, 2]. Long term, AKI can result in permanent kidney damage and chronic kidney disease (**CKD**) [3–5]. AKI and CKD cost the U.S. healthcare system up to $40 billion annually in all patients [6]. Despite their severe short- and long-term effects, critically ill neonates at high risk of AKI often go unrecognized due to the lack of neonatal AKI prediction models and risk stratification tools. Unidentified AKI and subsequent AKI-to-CKD can result in growth restriction, hypertension, increased cardiovascular risk, and kidney failure in children [4]. Early neonatal AKI risk identification could enable providers to intervene on modifiable factors to prevent kidney injury, CKD, and further complications [7].

Currently, the Baby Nephrotoxic Injury Negated by Just-in-Time Action (**NINJA**) is the only tool for identifying neonatal AKI risk [8]. However, Baby NINJA does not predict AKI occurrence and focuses solely on nephrotoxic medication-induced AKI, excluding other AKI etiologies and corresponding predisposing factors. This scope results in the tool missing a substantial number of high-risk neonates who develop AKI without prior nephrotoxic exposure. Similarly, the renal angina index (**RAI**) is a pediatric AKI risk assessment score designed to predict severe AKI within 48 to 72 hours of ICU admission in children but not neonates [9]. These knowledge gaps indicate a pressing need for more comprehensive and adaptable tools to predict and manage AKI risk in neonates effectively.

Monitoring SCr in critically ill neonates is challenging due to smaller blood volume, frequent needle sticks, and the need for central access, which can be distressing. A reliable AKI prediction tool would allow clinicians to reserve SCr monitoring for high-risk infants, minimizing unnecessary procedures and enabling timely, targeted care.

Predictive machine learning models help clinicians identify adult [10–12] and pediatric patients [13–15] at risk for AKI, but these tools are not yet available for the unique population of critically ill neonates. To address this significant clinical need, this study aimed to utilize novel machine-learning algorithms combined with readily available data in the electronic health records (**EHR**) to develop and validate a machine-learning model for neonatal AKI risk stratification.

## METHODS

### Study Design and Setting

This retrospective cohort study encompasses two comprehensive datasets from infants admitted to the main campus NICU at Nationwide Children’s Hospital (**NCH**). This NICU is one of the largest in the United States, with 130 beds and a diverse patient population and pathology, including two acute care units, one bronchopulmonary dysplasia unit, and a NICU stepdown unit. The average daily census in the main campus NICU is 60-80 neonates. The goal of the study was to develop and validate <u>NE</u>onatal <u>P</u>rotection of <u>H</u>ealth-<u>R</u>elated <u>O</u>utcomes (**NEPHRO**), a machine learning-based neonatal AKI prediction model.

#### Ethical Approval

The Institutional Review Board at NCH exempted this study due to its retrospective nature and was granted waiver of consent.

### Study Population

#### Inclusion and Exclusion Criteria

Patients were included if they were admitted to the acute care units at the NCH Main Campus NICU. Infants who were admitted to the bronchopulmonary dysplasia or the NICU stepdown units were excluded from the analysis due to different unit clinical and laboratory monitoring protocols. In addition, infants were excluded if UOP was recorded for less than 24 hours or if they had less than two SCr measurements.

The study included neonates who were admitted between 1/1/2017 and 6/30/2024, collecting data from the entire NICU stay, totaling n=8,059 patients and 206,220 patient days, achieving a representative range of neonatal pathology. We split our cohort into two non-overlapping datasets, one for training and cross-validation (patients admitted from 1/1/2017 to 12/31/2021, n= 5,443, 68% of total patients, 141,675 patient days), and one for temporal validation (patients admitted 01/01/2022 to 6/30/2024, n = 2,616, 32% of total patients = 64,545 patient days).

### Data Sources and Extraction

Data were extracted automatically from our EHR (EPIC, Clarity Data Warehouse) using structured query language.

After completing the automated data extraction, a nephrologist conducted a thorough manual review of randomly selected EHR charts (3% of the cohort) to ensure the accuracy and reliability of the automatically abstracted data. The nephrologist validated identifying neonates diagnosed with AKI to confirm that these patients were accurately captured. Next, they meticulously examined intake and output flowsheets to verify the precision of UOP documentation included in the dataset. Additionally, the nephrologist reviewed laboratory results to confirm the SCr trends, ensuring that the data accurately reflected these trends. This manual validation process confirmed the accuracy of the variables extracted automatically and included in the dataset, thereby ensuring that the model developed and validated subsequently was based on reliable and precise data.

### Variable Definitions

#### Outcome definition

Key variables included the outcome of neonatal AKI defined using the neonatal-modified KDIGO criteria as either a rise in serum creatinine ≥0.3 mg/dL or ≥50% from the lowest prior value, or urine output <1 mL/kg/h for 24 hours, consistent with definitions used in prior neonatal studies, [1]. In patients who experienced more than one episode of AKI, each episode was predicted separately after SCr and/or UOP returned to baseline for at least 24 hours, and patients met the AKI definition criteria again.

#### Outcome Labeling for Model Training

For model training and validation, we used all observations from patients who had no episodes of AKI as negative exemplars and only observations from 0-48 hours before the onset of an episode of AKI as positives. We chose 0–48-hour prediction window to allow time for implementing actions that could mitigate AKI or its progression. The remaining observations from patients with AKI (i.e., > 48 hours before AKI, during AKI, after AKI) were intentionally left out. This process ensured clearly separating positive and negative observations.

#### Neonatal AKI Risk Factors

A review of the literature and expert opinions identified risk factors for neonatal AKI, encompassing twenty-seven potential predisposing factors [16–24]. We collected demographic information, fluid balance, comorbid diagnoses, medications, laboratory results, surgical procedures, and the level of respiratory support. A list of the included risk factors and data dictionary is presented in **Supplemental Files 1** and **2**, respectively.

#### Missing Data

The frequency of SCr measurements differed among various patient encounters (**Supplemental File 3**).SCr criteria defined AKI only when more than two SCr measurements were available for analysis. UOP monitoring was standardized in the two acute care units. Diaper weights and/or volume of urine in catheterized patients were recorded every three hours and available for all infants throughout their NICU stay.

### Time-Varying Dataset Construction

Apart from patient demographics, all other variables were collected longitudinally throughout the NICU stay. Predictive comorbidities were abstracted from the EHR using ICD-9 and ICD-10 diagnostic codes. Significant nephrotoxic exposure was defined as treatment with nephrotoxic medication in the preceding 48 hours, using an established list consisting of 57 medications per the Baby NINJA criteria [8] (**Supplemental File 4**). Fluid balance was calculated using the gold standard method, which is the percent change in body weight [25, 26]. Congenital anomalies of the kidneys and urinary tract (**CAKUT**) were classified as mild/moderate and severe depending on how they affected nephron endowment and/or bladder function, and the diagnoses were abstracted using ICD codes (**Supplemental File 5**). Surgical procedures were classified as either minor or major based on their anticipated complexity, duration, and anesthesia requirements (**Supplemental File 6**).

#### Dataset Description

We describe our dataset in terms of all collected features by reducing variables to the patient level. We compared variables between the training set and the temporal validation set to evaluate similarities and potential differences.

### Univariate Analysis

We conducted a univariate logistic regression analysis to study the relationship between 27 patient characteristics and the odd ratios of developing AKI. Binary variables are expressed by the total number and percentages of patients affected and were compared by using Fisher’s exact test. Continuous variables are presented as median and interquartile range (IQR) and were compared by using the Wilcoxon rank-sum test. Employing a Bonferroni correction to account for multiple comparisons, a p-value < 0.0017 was considered statistically significant.

### Modelling

#### Time Dependency

While some of our variables are static (e.g., patient demographics), most variables are time-dependent to reflect the real-time changes in AKI risk during the NICU encounter. We built a time-grid dataset where each variable is evaluated every 12 hours, once at 9 AM and once at 9 PM, for every patient encounter, resulting in 412,440 total data points/observations of each of the 27 time-dependent variables. We chose those time points (9 AM and 9 PM) in accordance with the clinical rounding time for NICU providers to make the prediction timely and clinically relevant. The model evaluate variables every 12 hours and predicts AKI in the next 0-48 hours before changes in Scr and UOP occur, **Figure 1**.

**Figure 1:**
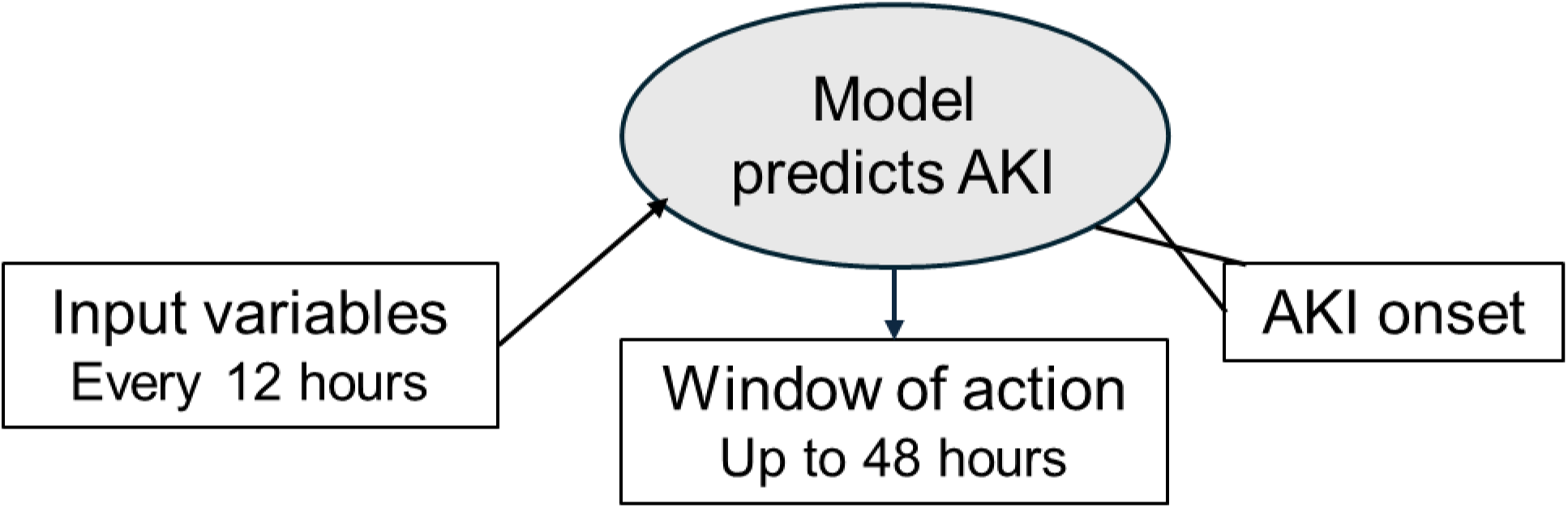
Time Dependent Variables, AKI Prediction and Window of Action in Relation to AKI Onset. Timing of AKI prediction relative to conventional biomarker-defined AKI onset. Prediction creates a 48-hour window during which real-time risk scores enable early identification of high-risk neonates to allow timely interventions that can prevent kidney injury.

#### Machine Learning Model

We chose logistic regression with Least Absolute Shrinkage and Selection Operator (**LASSO**) regularization as our machine-learning model development method. LASSO is a linear model (in log-odds space), therefore providing transparent and interpretable predictions where each variable either increases, decreases, or does not affect the predicted outcome [27]. The LASSO regularization uses a penalty on the absolute regression coefficients to perform variable selection, shrinking some coefficients to zero. This process leaves only the most significant neonatal AKI predictors, simplifying high-dimensional datasets to a manageable and actionable subsets [27]. LASSO’s transparency allows understanding of an individual feature’s contribution to neonatal AKI risk, which is critical for clinical decisions and lays the groundwork for further analysis, model comparison, and future research.

The LASSO model has one hyperparameter, the degree or regularization, which was chosen based on a patient-level 10-fold cross-validation by minimizing mean-squared error [27]. LASSO optimization was implemented in Python using the scikit-learn library. During optimization, each observation was weighted by one over the total number of observations per patient to control for different lengths of stays across patients.

#### Machine Learning Validation and Evaluation

We evaluated the performance of the LASSO model using the area under the receiver operating characteristic (**AUROC**) curve and precision-Recall curves (F1-score). The AUROC compares sensitivity (true positive rate) and specificity (one minus false positive rate) for all possible decision thresholds of the model [28]. F1 score balances the trade-off between precision and recall and is useful when predicting rare events [29]. We validate the model’s generalizability by computing the AUROC curve on the temporal validation set, which was not used during training. For comparison, we also compute the cross-validated AUROC curve for the training set. AUROC curves and areas are computed based on weighted observations (one over the number of observations per patient). We provide 95% confidence intervals for all reported metrics that are computed based on bootstrapping observations (sampling with replacement at the patient level with 100 repetitions).

#### Real-Time Clinical Simulation

We also investigated the LASSO model’s alarm rate and sensitivity in a real-time clinical simulation. In the simulation, the model is applied twice per day for the entire encounter of all patients in the temporal validation cohort. For all potential intervention risk thresholds, we compute the alarm rate in terms of the average percentage of patients that are flagged as high risk for developing AKI and compare it to sensitivity, the percentage of AKI episodes that would be predicted within 48 hours before clinically detectable changes in UOP and/or SCr. We also compute the positive predictive value (**PPV**) at the prediction level and the PPV lift (change from baseline predictability).

## RESULTS

### Training and Temporal Validation Datasets

Patient characteristics for the training and validation cohorts are given in **Table 1**. The prevalence of certain variables was similar between the training and validation datasets. However, other variables revealed significant differences between the two datasets. For instance, age at admission to the NICU, comorbid diagnoses such as necrotizing enterocolitis, major surgeries, respiratory support, and exposure to nephrotoxic medications varied significantly (**Table 1)**.

**Table 1:** Patient and Dataset Characteristics for the Training and Validation Cohorts.

| Variables | Derivation/Training<br>(N = 5,443) | Temporal Validation<br>(N = 2,616) | p-value |
| --- | --- | --- | --- |
| **Demographic** |  |  |  |
| Gestational age (**GA**) in weeks (range) | 36.7 (32.0, 39.0) | 36.4 (31.9, 38.7) | 1.000 |
| GA < 28 weeks, n (%) | 665 (12.2) | 348 (13.3) | 0.173 |
| GA 28-31 weeks, n (%) | 525 (9.6) | 243 (9.3) | 0.627 |
| GA 32-37 weeks, n (%) | 1,802 (33.1) | 850 (32.5) | 0.595 |
| GA > 37 weeks, n (%) | 2,448 (45.0) | 1,173 (44.8) | 0.924 |
| Age at admission in days (range) | 2.8 (0.7, 9.8) | 2.1 (0.7, 9.1) | <0.001* |
| Sex (male), n (%) | 3,074 (56.5) | 1,446 (55.3) | 0.314 |
| **Comorbidities, n (%)** |  |  |  |
| CAKUT (mild or moderate) | 80 (1.5) | 40 (1.5) | 0.845 |
| CAKUT (severe) | 21 (0.4) | 18 (0.7) | 0.085 |
| CAKUT (vesicoureteral reflux) | 23 (0.4) | 14 (0.5) | 0.485 |
| Inborn errors of metabolism | 287 (5.3) | 199 (7.6) | <0.001* |
| Congenital heart disease | 769 (14.1) | 395 (15.1) | 0.250 |
| Structural disorder of the heart | 998 (18.3) | 510 (19.5) | 0.211 |
| Patent Ductus Arteriosus (**PDA**) | 678 (12.5) | 346 (13.2) | 0.335 |
| Necrotizing Enterocolitis (**NEC**), Stage 2+ | 230 (4.2) | 154 (5.9) | 0.001* |
| **Surgeries, n (%)** |  |  |  |
| Had surgery (any risk) | 1,099 (20.2) | 492 (18.8) | 0.151 |
| Had surgery (mild risk) | 742 (13.6) | 341 (13.0) | 0.485 |
| Had surgery (greater risk) | 520 (9.6) | 212 (8.1) | 0.035 |
| **Respiratory support, n (%)** |  |  |  |
| On respiratory support | 2,646 (48.6) | 1,445 (55.2) | <0.001* |
| On non-invasive respiratory support | 1,949 (35.8) | 1,148 (43.9) | <0.001* |
| On invasive respiratory support | 1,683 (30.9) | 817 (31.2) | 0.777 |
| Avg. % fraction inspired O<sub>2</sub> (range) | 26.6 (22.2, 34.6) | 23.7 (21.4, 28.5) | <0.001* |
| **Medication administration, n (%)** |  |  |  |
| Nephrotoxic medications | 2,370 (43.5) | 1,058 (40.4) | 0.009 |

| Variables | Deriva-<br>tion/Training<br>(N = 5,443) | Temporal Valida-<br>tion<br>(N = 2,616) | p-value |
| --- | --- | --- | --- |
| IV Vasopressor | 215 (4.0) | 71 (2.7) | 0.005 |
| Contrast Ioversol | 757 (13.9) | 285 (10.9) | 0.001* |
| **Labs, n (%)** |  |  |  |
| Blood culture | 1,634 (30.0) | 939 (35.9) | <0.001* |
| Serum Creatinine | 4,572 (84.0) | 2,191 (83.8) | 0.796 |
| **Urine Output** |  |  |  |
| Avg. 24h-UOP in mL/kg/hr | 4.0 (3.5, 4.4) | 4.0 (3.5, 4.4) | 0.822 |
| Avg. Fluid Balance | 0.02 (-0.02, 0.04) | 0.01 (-0.02, 0.04) | 0.564 |

**CAKUT:** Congenital anomalies of the kidneys and urinary tract. **UOP:** urinary output. All variables are computed at the patient level. Data are represented as proportion and percentage (binary variables) or median and interquartile range (continuous variables). P-values are based on Fisher's exact test for binary variables and Wilcoxon rank-sum test for continuous variables. Asterisks (\*) denote statistical significance.

### Prevalence of Neonatal AKI

Across the training and validation datasets and using nm-KDIGO, 881 neonates (11% of patients) developed AKI during the NICU stay. Among patients with AKI, 83.8% experienced a single episode, 10.9% had two distinct episodes, and the remaining 5.3% had more than two episodes, resulting in a total of 1,127 AKI episodes. Out of all those AKI episodes, 440 (39%) were initiated by meeting the SCr increase criteria, and 687 (61%) met the UOP reduction criteria.

We compared the prevalence of all variables among patients with and without AKI and found significant differences in AKI risk factors between the two groups (**Table 2)**. This comparison highlights the distinct profiles of neonates at risk for AKI.

**Table 2:** Patient Characteristics for the Acute Kidney Injury (AKI) and non-AKI Cohorts.

| Variables | Patients with AKI<br>(N = 881) | Patients without<br>AKI<br>(N = 7,178) | p-value |
| --- | --- | --- | --- |
| **Demographic** |  |  |  |
| Gestational age (**GA**) in weeks (range) | 34.9 (28.1, 38.3) | 36.9 (32.4, 39.0) | 1.000 |
| GA < 28 weeks, n (%) | 208 (23.6) | 805 (11.2) | p<0.001 |
| GA 28-31 weeks, n (%) | 104 (11.8) | 664 (9.3) | 0.018 |
| GA 32-37 weeks, n (%) | 253 (28.7) | 2,399 (33.4) | 0.005 |
| GA > 37 weeks, n (%) | 314 (35.6) | 3307 (46.1) | p<0.001* |
| Age at admission in days (range) | 2.1 (0.7, 12.1) | 2.8 (0.7, 9.1) | 0.008 |
| Sex (male), n (%) | 525 (59.6) | 3,995 (55.7) | 0.028 |
| **Comorbidities, n (%)** |  |  |  |
| CAKUT (mild or moderate) | 36 (4.1) | 84 (1.2) | p<0.001* |
| CAKUT (severe) | 16 (1.8) | 23 (0.3) | p<0.001* |
| CAKUT (vesicoureteral reflux) | 11 (1.2) | 26 (0.4) | 0.001* |
| Inborn errors of metabolism | 119 (13.5) | 367 (5.1) | p<0.001* |
| Congenital heart disease | 201 (22.8) | 963 (13.4) | p<0.001* |
| Structural disorder of the heart | 215 (29.9) | 1,293 (17.6) | p<0.001* |
| Patent Ductus Arteriosus (PDA) | 220 (25.0) | 804 (11.2) | p<0.001* |
| Necrotizing Enterocolitis (NEC), Stage 2+ | 96 (10.9) | 288 (4.0) | p<0.001* |
| **Surgeries, n (%)** |  |  |  |
| Had surgery (any risk) | 391 (44.4) | 1,200 (16.7) | p<0.001* |
| Had surgery (mild risk) | 265 (30.1) | 818 (11.4) | p<0.001* |
| Had surgery (greater risk) | 214 (24.3) | 518 (7.2) | p<0.001* |
| **Respiratory support, n (%)** |  |  |  |
| On respiratory support | 626 (71.1) | 3,465 (48.3) | p<0.001* |
| On non-invasive respiratory support | 395 (44.8) | 2,702 (37.6) | p<0.001* |
| On invasive respiratory support | 566 (64.2) | 1,934 (26.9) | p<0.001* |
| Avg. % fraction inspired O<sub>2</sub> (range) | 29.5 (24.0, 39.5) | 24.6 (21.4, 30.5) | p<0.001* |
| **Medication administration, n (%)** |  |  |  |
| Nephrotoxic medications | 536 (60.8) | 2,892 (40.3) | p<0.001* |
| IV Vasopressor | 140 (15.9) | 146 (2.0) | p<0.001* |
| Contrast Ioversol | 224 (25.4) | 818 (11.4) | p<0.001* |
| **Labs, n (%)** |  |  |  |
| Blood culture | 481 (54.6) | 2,092 (29.1) | p<0.001* |
| Serum Creatinine | 808 (91.7) | 5,955 (83.0) | p<0.001* |
| **Urine Output** |  |  |  |
| Avg. 24h-UOP in mL/kg/hr | 3.7 (3.1, 4.1) | 4.0 (3.6, 4.5) | p<0.001* |
| Avg. Fluid Balance | 0.04 (0.01, 0.06) | 0.01 (-0.02, 0.04) | p<0.001* |

**CAKUT:** Congenital anomalies of the kidneys and urinary tract. **UOP:** urinary output.

### Univariate predictors of neonatal AKI

The univariate logistic regression analysis revealed several key risk factors for neonatal AKI (**Table 3**). Lower gestational age, reduced 24-hour UOP, increased fluid balance, and elevated SCr, even when not meeting neonatal AKI KDIGO criteria, significantly increased AKI risk. Invasive ventilatory support and higher fractional-inspired oxygen levels were significant AKI predictors.

**Table 3:**
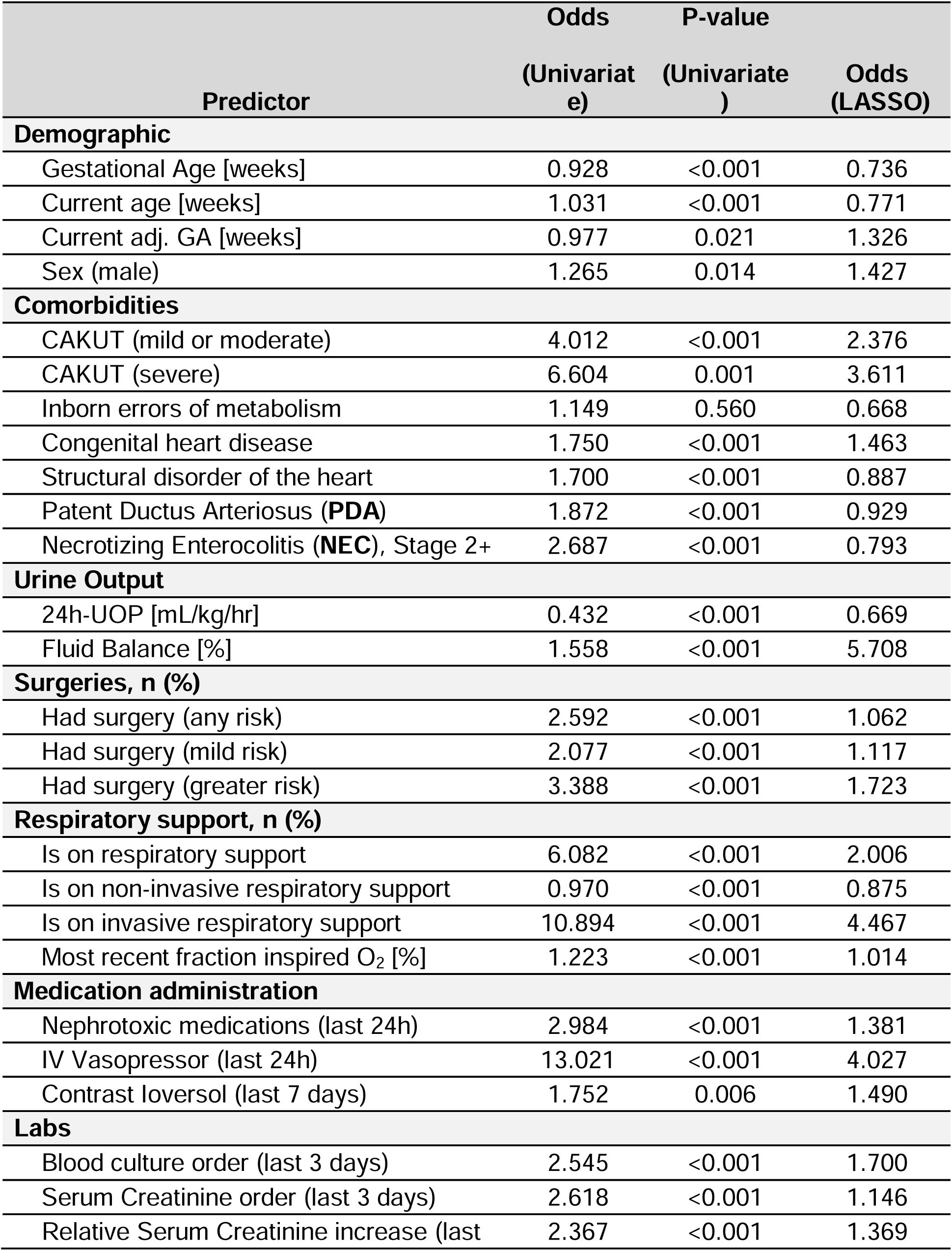
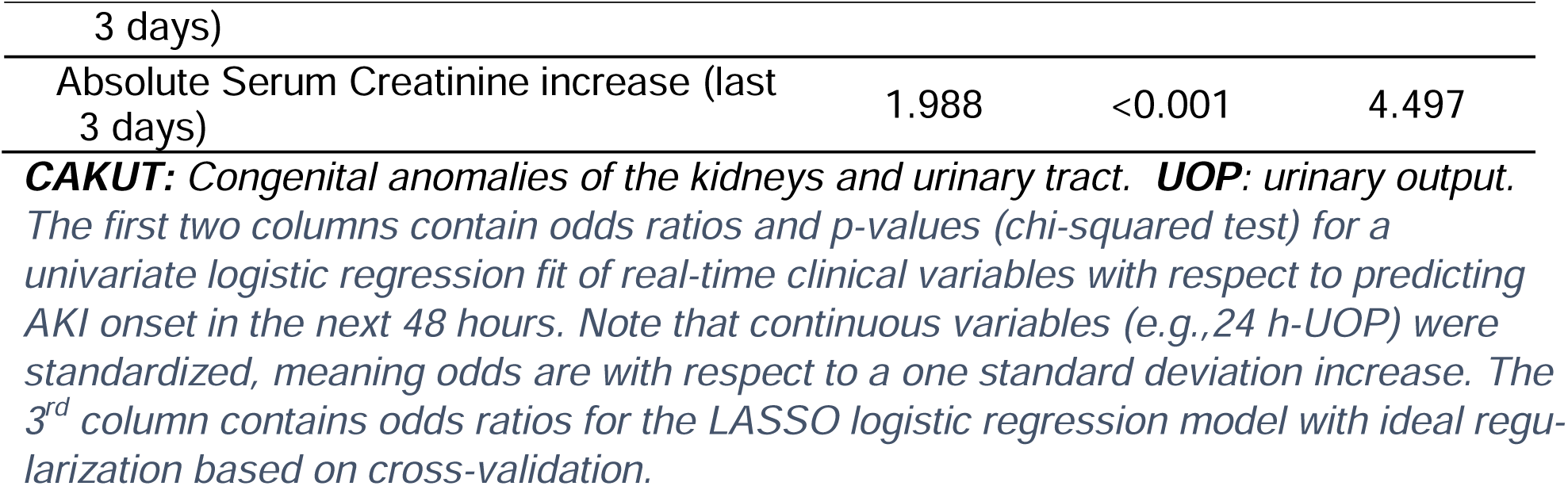
Comparison of Univariate and LASSO Logistic Regression for Predicting Neonatal Acute Kidney Injury (AKI)

Medications, namely intravenous vasopressors and nephrotoxic medications, surgical interventions, recent laboratory orders for blood cultures, and frequent SCr monitoring were significant predictors of AKI. Both mild/moderate and severe congenital anomalies of the kidney and urinary tract (**CAKUT**) correlated with higher AKI odds, with severe CAKUT showing the most significant risk. Major surgical procedures also substantially elevated the risk of AKI (**Table 3**).

### Nephrotoxic Medications-Associated Neonatal AKI

Our data indicate that 69.6% of neonatal AKI episodes (784 of 1,127) occurred without prior significant nephrotoxic medication exposure in the 72 hours preceding neonatal AKI development (**Figure 2)**. This observation indicates that other predisposing factors also have a substantial role in neonatal AKI.

**Figure 2:**
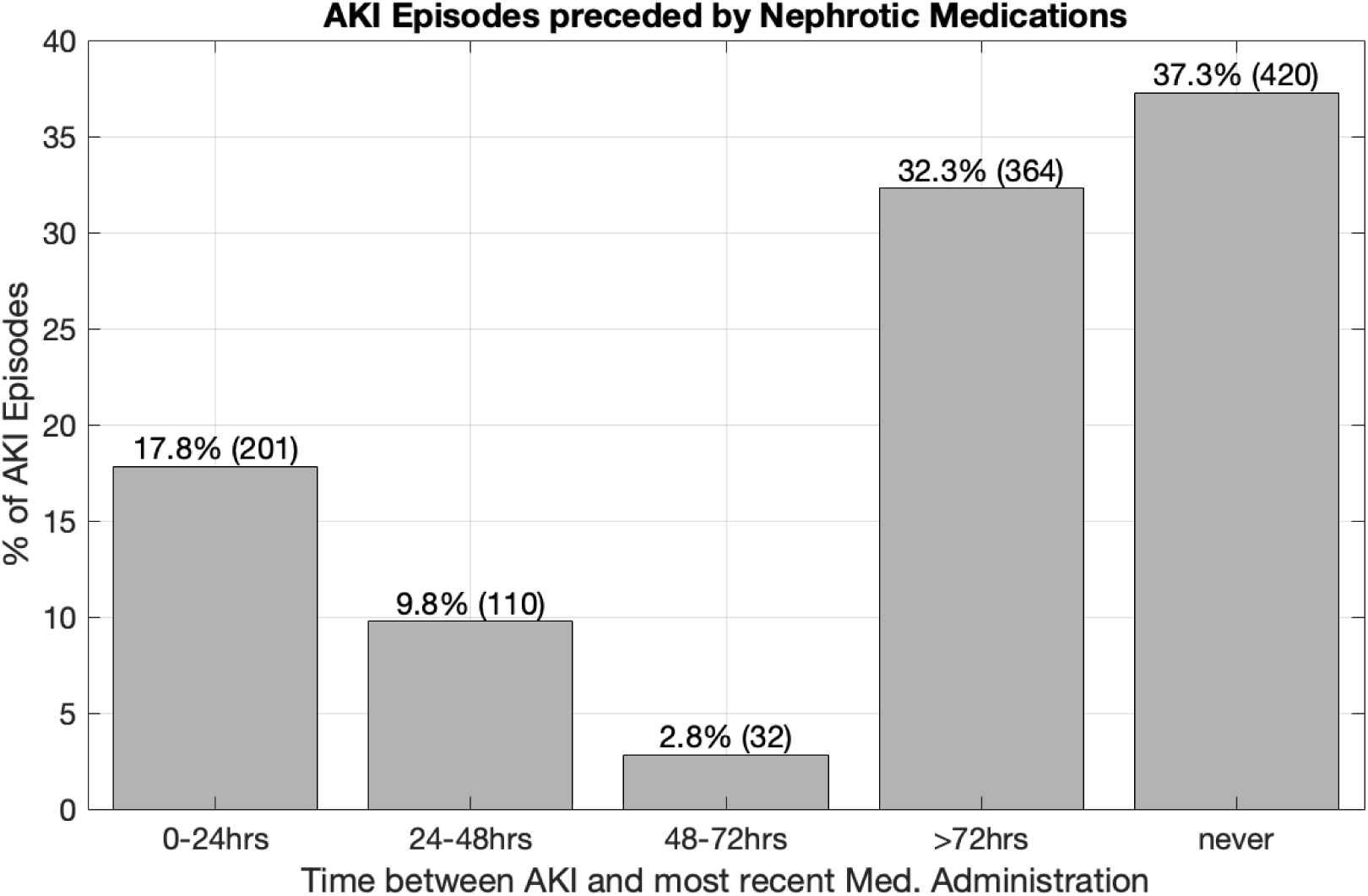
Neonatal AKI Episodes Preceded by Nephrotoxic Medications. Percentage and counts of AKI episodes that were preceded by a significant nephrotoxic medication administration up to 72 hours before the onset of AKI, compared to episodes that were not preceded by a significant nephrotoxic medication administration.

### Neonatal AKI Prediction

#### LASSO Model

The LASSO model with ideal regularization (based on minimum cross-validation error) kept all 27 variables (**Table 3**). The model predicts neonatal AKI 0-48 hours before significant changes in SCr and UOP occur, with a cross-validated area under the AUROC curve of 0.814 (95% CI: 0.787, 0.843) **(Figure 3**). The model’s predictive performance generalized to the temporal validation cohort, with an AUROC of 0.815 (0.795, 0.834).

**Figure 3:**
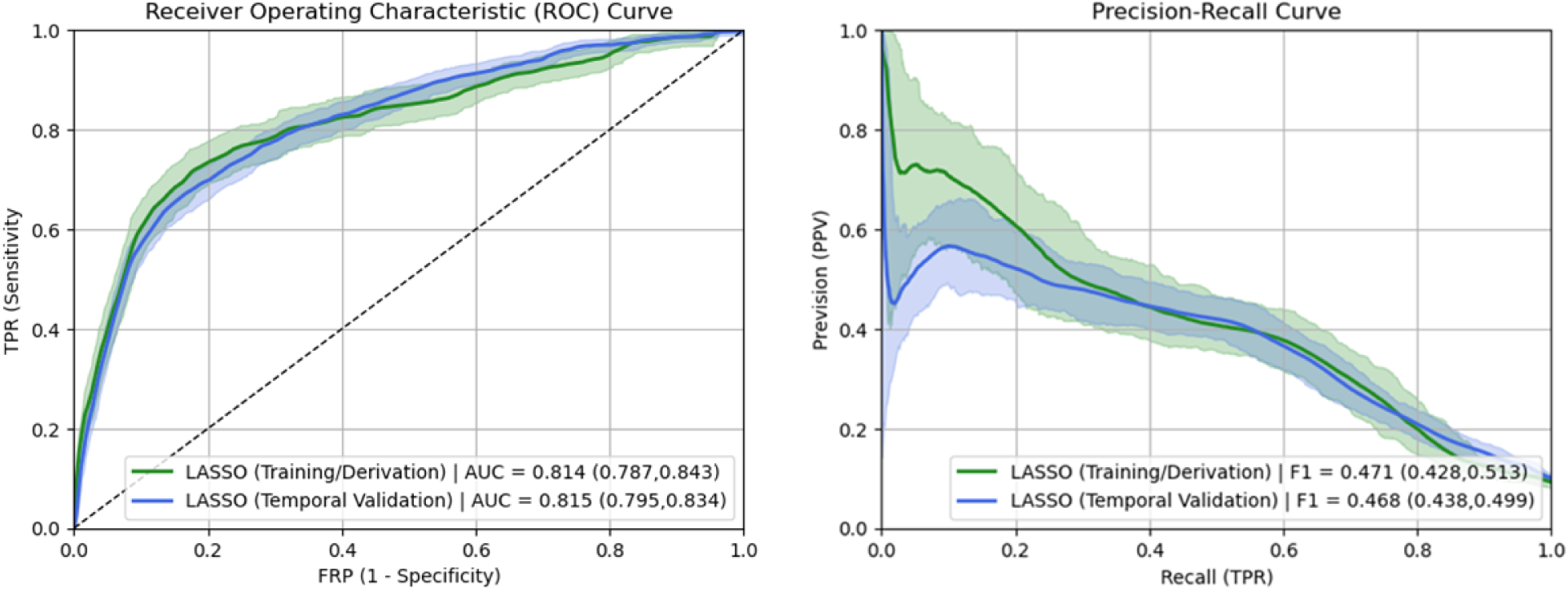
The Performance of Neonatal AKI Predictive Model. Left: Patient-weighted ROC curve and area-under-the-curve (AUC) of the LASSO model. Right: Patient-weighted precision-recall curve of the LASSO model. The estimate on the training set is based on a cross-validated estimate. Brackets show 95% confidence intervals.

#### Real-time Clinical Model Simulation

Our model relies on routinely collected risk factors that are readily available in the EHR. We simulated clinical implementation of the LASSO model, computing AKI risk at 9 AM and 9 PM throughout the NICU hospitalizations for all 2,616 patients in our temporal validation cohort. Model sensitivity (in terms of % of AKI events predicted 0-48 hours before they occur), for all possible intervention thresholds, is illustrated in **Figure 4A**. The positive predictive values are given in **Figure 4B**.

**Figure 4:**
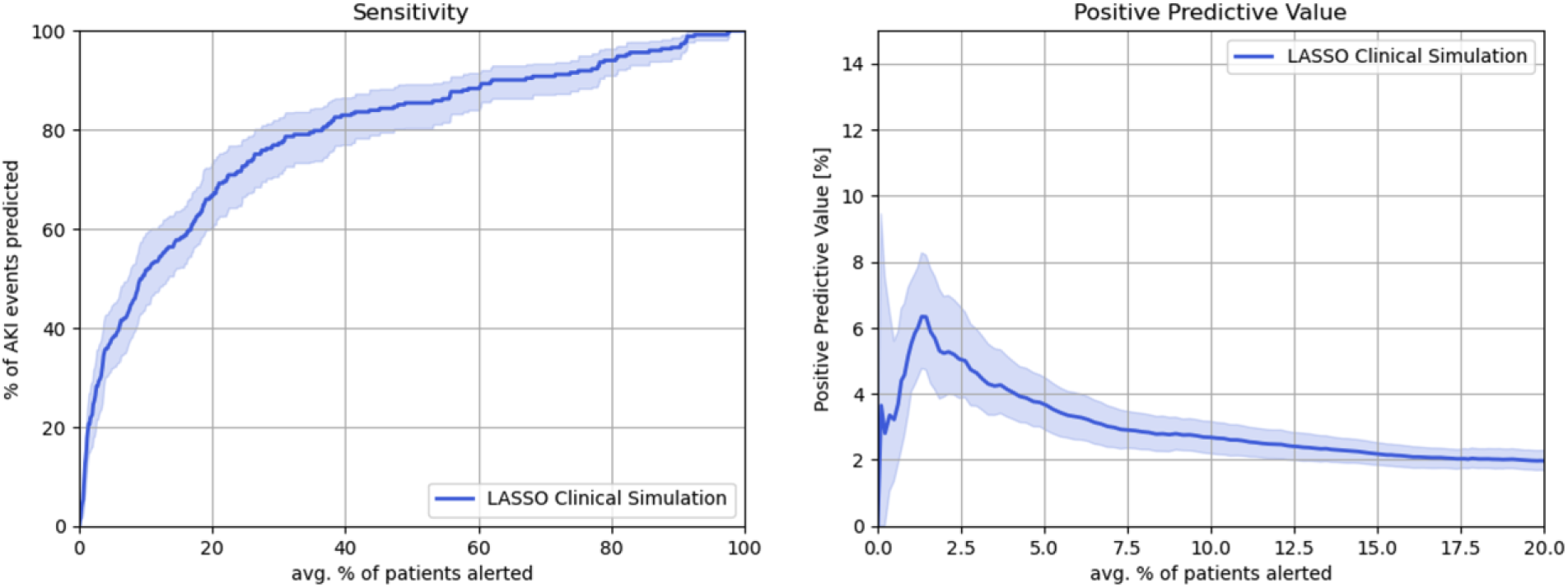
Results of Clinically Simulating a LASSO Model in Real-Time for Predicting Neonatal AKI. Results of clinical simulating the LASSO model in real-time. The X-axis shows the average % of patients that are alerted when risk is evaluated for all possible alert thresholds. Y-axis (left) shows the % of AKI events that an alert would predict between 0-48 hrs ahead of time. Y-axis (right) shows the positive predictive value of the model.

We established two thresholds for identifying neonatal AKI risk: one indicating a high probability and the other a very high probability of developing neonatal AKI (**Supplemental File 7**). These thresholds classify 15% and 5% of all admitted patients as high risk and very high risk, respectively. On average, these criteria will identify eight patients per day (ranging from 5-20) as high risk and four patients per day (ranging from 0-8) as very high risk. A high-probability patient under the optimal LASSO model is 3.2 times more likely to have AKI in the next 48 hours than the average NICU patient, and a very high-probability patient is 5.4 times more likely (**Supplemental File 7**).

## DISCUSSION

This study aims to overcome current limitations and address a huge clinical need for the vulnerable, critically ill neonatal population. We utilized a novel and comprehensive approach to develop <u>NE</u>onatal <u>P</u>rotection of <u>H</u>ealth-<u>R</u>elated <u>O</u>utcomes (**NEPHRO**), an automated machine learning-based neonatal AKI prediction model integrated into the EHR to assist clinicians with identifying neonates at high risk of developing AKI. Our approach uses a comprehensive list of AKI risk factors coupled with the power of LASSO that ensures complete transparency and interpretability [27]. We utilized two comprehensive datasets for model training and temporal validation, which exhibited similarities in certain variables and differences in others. Despite these differences in variable prevalence, the model’s performance was comparable across both datasets, demonstrating its adaptability and potential generalizability across different settings. Using a large single-center cohort of critically ill neonates and the known and potential neonatal AKI risk factors, our study demonstrates that applying machine learning algorithms combined with EHR data can accurately predict neonatal AKI.

To our knowledge, this risk stratification tool is the first known machine-learning-based neonatal AKI prediction model. Machine learning models are effective in predicting AKI and AKI outcomes, as disclosed in the studies by *Sandokjj et al*. for pediatric AKI [15], *Dong et al*. for critically ill pediatric patients [13], and *Deng et al*. for predicting pediatric AKI major adverse kidney outcomes [30]. The AUROC assessed the performance of those models and was 0.76 using LASSO, 0.89 using the age-dependent ensemble model, and 0.75-0.81 using LASSO or extremely boosted gradient, respectively. Of note, *Deng et al*. reported higher AUC with extremely boosted gradient compared to LASSO [30].

Predictive machine learning tools help clinicians identify adult and pediatric patients at risk for AKI [10–15], allowing time to intervene before kidney injury occurs. Still, these tools are not yet available for neonates [7]. Baby NINJA is the only currently available initiative to prevent neonatal AKI [8]. Our study demonstrated that pathologic fluid balance (fluid overload) and treatment with invasive ventilation and vasopressors are associated with higher odds of neonatal AKI development compared to nephrotoxic medications. These findings suggest this prediction model, which includes other relevant AKI predictors, may augment existing tools like Baby NINJA by capturing broader risk pathways.

Traditional neonatal AKI risk scores rely on manual data and lack real-time updates, making them inefficient and less reliable [31]. While the RAI predicts AKI in older children, it is not practical for neonates with prolonged hospital stays, [9]. Our model is automated, updated and accurately predicts AKI up to 48 hours before SCr or UOP changes, enabling early intervention and improved outcomes.

This study represents a pioneering advancement in utilizing innovative machine-learning techniques for interventions in neonatal kidney diseases, with a specific focus on AKI in critically ill neonates. Our model integrates with the EHR to provide continuously updated, real-time risk scores throughout NICU hospitalization, allowing early identification of high-risk neonates. This enables timely, targeted interventions on modifiable risk factors, improves risk stratification, and supports more effective clinical decision-making and resource use.

The strengths of our study include developing a novel and comprehensive, evidence-based neonatal AKI prediction model leveraging machine learning algorithms to address a critical clinical need within the neonatal population. This innovative model is longitudinally designed, seamlessly integrated with the EHR, and presented as a real-time risk stratification score that updates hourly throughout NICU hospitalization. Such a dynamic and integrated approach to neonatal kidney disease has not been previously reported, marking a significant advancement in the field and setting a new standard for predictive healthcare in neonatal nephrology.

Our study has limitations. First, the model is developed and trained on a dataset from a single center-level IV NICU. Although one of the largest NICUs in the United States, these findings need to be validated externally in subsequent studies and also in lower acuity NICU settings. This limitation will be addressed in future studies. Second, we only used LASSO to develop the machine learning algorithm due to its known transparency and interpretability. Previous studies in pediatric AKI established that extremely boosted gradient models might outperform LASSO in accuracy [14]. Although neonatal AKI is a unique entity and different from pediatric AKI, our team is currently working on training various machine learning algorithms, such as extremely boosted gradient models and neural networks, to address this limitation. Third, the model requires further external testing and prospective validation in clinical trials for future scalable implementation. Lastly, the potential for development of alert fatigue from false positives should be evaluated in future studies.

In conclusion, to our knowledge, this real-time EHR-based model is the first to predict AKI risk in critically ill neonates using routinely collected data. This comprehensive, evidence-based tool is poised to revolutionize current neonatal AKI practices by empowering clinicians to intervene on modifiable risk factors before kidney injury occurs, thus preventing AKI and potentially improving both short- and long-term outcomes for critically ill neonates. Operating in real-time and fully automated, our model is designed for seamless integration with the EHR, promising to enhance accuracy and broaden its applicability across diverse clinical settings. This advancement has the potential to improve neonatal kidney care and patient outcomes on a large scale. Future studies will focus on externally validating the model and implementing it locally to evaluate performance in real-world practice.

## Data Availability

All data produced in the present study are available upon reasonable request to the authors

## REFERENCES

1. Jetton, J.G., et al., Incidence and outcomes of neonatal acute kidney injury (AWAKEN): a multicentre, multinational, observational cohort study. Lancet Child Adolesc Health, 2017. 1(3): p. 184–194.

2. Starr, M.C., et al., Advances in Neonatal Acute Kidney Injury. Pediatrics, 2021. 148(5).

3. Harer, M.W., et al., Follow-up of Acute kidney injury in Neonates during Childhood Years (FANCY): a prospective cohort study. Pediatr Nephrol, 2017. 32(6): p. 1067–1076.

4. Mammen, C., et al., Long-term risk of CKD in children surviving episodes of acute kidney injury in the intensive care unit: a prospective cohort study. Am J Kidney Dis, 2012. 59(4): p. 523–30.

5. Maqsood, S., et al., Outcome of extremely low birth weight infants with a history of neonatal acute kidney injury. Pediatr Nephrol, 2017. 32(6): p. 1035–1043.

6. Ozieh, M.N., et al., Trends in healthcare expenditure in United States adults with chronic kidney disease: 2002-2011. BMC Health Serv Res, 2017. 17(1): p. 368.

7. Mohamed, T., et al., Evidence-based risk stratification for neonatal acute kidney injury: a call to action. Pediatr Nephrol, 2025.

8. Stoops, C., et al., Baby NINJA (Nephrotoxic Injury Negated by Just-in-Time Action): Reduction of Nephrotoxic Medication-Associated Acute Kidney Injury in the Neonatal Intensive Care Unit. J Pediatr, 2019. 215: p. 223–228.e6.

9. Basu, R.K., et al., Derivation and validation of the renal angina index to improve the prediction of acute kidney injury in critically ill children. Kidney Int, 2014. 85(3): p. 659–67.

10. Ugwuowo, U., et al., Real-Time Prediction of Acute Kidney Injury in Hospitalized Adults: Implementation and Proof of Concept. Am J Kidney Dis, 2020. 76(6): p. 806–814.e1.

11. Yue, S., et al., Machine learning for the prediction of acute kidney injury in patients with sepsis. J Transl Med, 2022. 20(1): p. 215.

12. Zhang, Z., K.M. Ho, and Y. Hong, Machine learning for the prediction of volume responsiveness in patients with oliguric acute kidney injury in critical care. Crit Care, 2019. 23(1): p. 112.

13. Dong, J., et al., Machine learning model for early prediction of acute kidney injury (AKI) in pediatric critical care. Crit Care, 2021. 25(1): p. 288.

14. Luo, X.Q., et al., Machine Learning-Based Prediction of Acute Kidney Injury Following Pediatric Cardiac Surgery: Model Development and Validation Study. J Med Internet Res, 2023. 25: p. e41142.

15. Sandokji, I., et al., A Time-Updated, Parsimonious Model to Predict AKI in Hospitalized Children. J Am Soc Nephrol, 2020. 31(6): p. 1348–1357.

16. Cataldi, L., et al., Potential risk factors for the development of acute renal failure in preterm newborn infants: a case-control study. Arch Dis Child Fetal Neonatal Ed, 2005. 90(6): p. F514–9.

17. Charlton, J.R., et al., Incidence and Risk Factors of Early Onset Neonatal AKI. Clin J Am Soc Nephrol, 2019. 14(2): p. 184–195.

18. Fan, Y., et al., Risk factors and outcomes of acute kidney injury in ventilated newborns. Ren Fail, 2019. 41(1): p. 995–1000.

19. Garg, P.M., et al., Severe acute kidney injury in neonates with necrotizing enterocolitis: risk factors and outcomes. Pediatr Res, 2021. 90(3): p. 642–649.

20. Hu, Q., et al., Risk Factors for Acute Kidney Injury in Critically Ill Neonates: A Systematic Review and Meta-Analysis. Front Pediatr, 2021. 9: p. 666507.

21. Lee, J.H., et al., Risk factors of acute kidney injury in children after cardiac surgery. Acta Anaesthesiol Scand, 2018. 62(10): p. 1374–1382.

22. Li, S., et al., Incidence, risk factors, and outcomes of acute kidney injury after pediatric cardiac surgery: a prospective multicenter study. Crit Care Med, 2011. 39(6): p. 1493–9.

23. Mohamed, T., C. Mpody, and O. Nafiu, Perioperative Neonatal Acute Kidney Injury is Common: Risk Factors for Poor Outcomes. Am J Perinatol, 2023.

24. Perico, N., et al., Maternal and environmental risk factors for neonatal AKI and its long-term consequences. Nat Rev Nephrol, 2018. 14(11): p. 688–703.

25. Selewski, D.T., et al., The impact of fluid balance on outcomes in critically ill near-term/term neonates: a report from the AWAKEN study group. Pediatr Res, 2019. 85(1): p. 79–85.

26. Selewski, D.T., et al., The impact of fluid balance on outcomes in premature neonates: a report from the AWAKEN study group. Pediatr Res, 2020. 87(3): p. 550–557.

27. Tibshirani, R., Regression Shrinkage and Selection via the LASSO. 1995.

28. Mandrekar, J.N., Receiver operating characteristic curve in diagnostic test assessment. J Thorac Oncol, 2010. 5(9): p. 1315–6.

29. Boyd, K., Area under the Precision-Recall Curve: Point Estimates and Confidence Intervals, K.H.E.C.D. Page, Editor. 2013, Springer Nature.

30. Deng, Y.H., et al., Outcome prediction for acute kidney injury among hospitalized children via eXtreme Gradient Boosting algorithm. Sci Rep, 2022. 12(1): p. 8956.

31. Sethi, S.K., et al., Validation of the STARZ neonatal acute kidney injury risk stratification score. Pediatr Nephrol, 2022. 37(8): p. 1923–1932.

