## Supplemental Files for "Leveraging Machine Learning for Developing and Validating a Neonatal Acute Kidney Injury Prediction Model (NEPHRO): A Comprehensive Evidence-Based Neonatal AKI Risk Stratification Tool"

**Supplemental File #1:** AKI Risk Factors Based on Published Literature

| **Risk factor** | **Notes** |
|---|---|
| Out-born delivery | All our patients are considered out-born because we are not a birth hospital. |
| Resuscitation with epinephrine | |
| Hyperbilirubinemia | |
| Inborn errors of metabolism | |
| Surgical procedures | |
| Intubation | |
| Oligo/polyhydramnios | |
| Kidney anomalies | |
| Congenital heart disease | |
| NEC | |
| Surgical procedures | |
| Exposures | (vasopressors, NSAID) |
| Discharge diagnoses | PDA, NEC, Sepsis |

**Supplemental File #2:** Frequency of Serum Creatinine Monitoring.

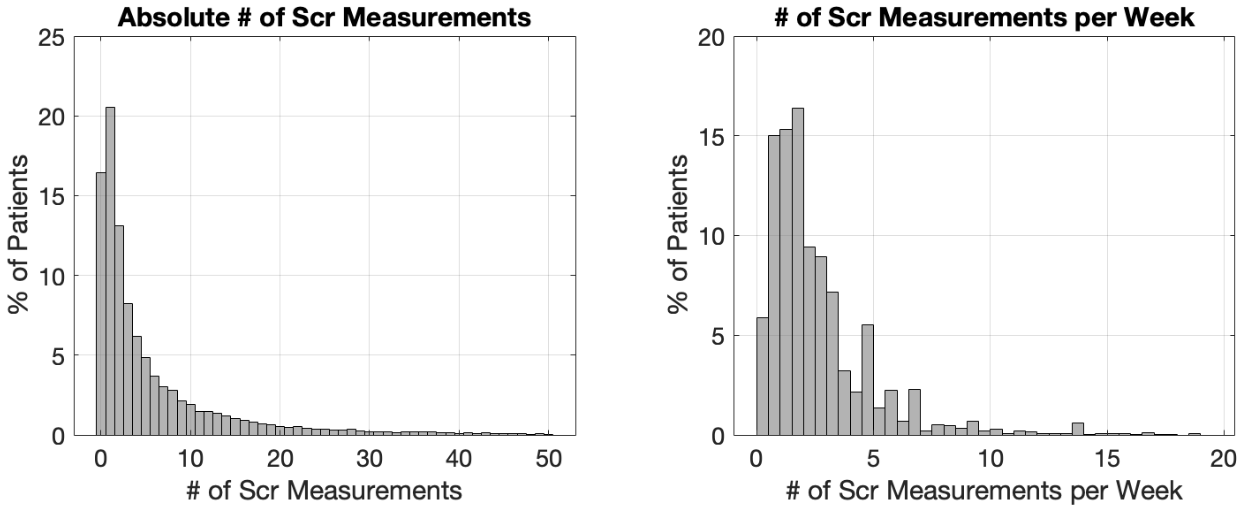


*Left: The absolute number of Scr measurements per NICU encounter. 16.4% of patients had no Scr labs and 20.5% of patients had only 1 Scr measurement. Right: The number of Scr measurements per week. Numbers are based on the entire 2017-2024 cohort.*

**Supplemental File #4: List of Nephrotoxic Medications (Stoops et al)**

List of nephrotoxic medications

| Acyclovir | Enalaprilat | Mesalamine |
|---|---|---|
| Ambisome[*] | Foscarnet | Methotrexate |
| Amikacin | Gadopentetate dimeglumine[*] | Nafcillin |
| Amphotericin B | Gadoextate disodium[*] | Piperacillin/Tazobactam |
| Captopril | Ganciclovir | Piperacillin |
| Carboplatin | Gentamicin | Sirolimus |
| Cefotaxime | Ibuprofen | Sulfasalazine |
| Ceftazidime | Ifosfamide | Tacrolimus |
| Cefuroxime | Iodixanol[*] | Ticarcillin/Clavulanic Acid |
| Cidofovir[*] | Iohexol[*] | Tobramycin |
| Cisplatin | Iopamidol[*] | Topiramate |
| Colistimethate | Ioversol[*] | Valacyclovir |
| Cyclosporine | Ketorolac | Valganciclovir |
| Dapsone | Lisinopril | Vancomycin |
| Enalapril | Lithium | Zonisamide |

**Supplemental File #5: congenital anomalies of the kidney and urinary tract classification**

| **Mild CAKUT** | **Moderate CAKUT** | **Severe CAKUT** |
|---|---|---|
| Hydronephrosis | Vesicoureteric reflux grade III | Posterior urethral valves |
| Hydroureter | Vesicoureteric relux grade IV | Prune Belly Syndrome |
| Unilateral renal agenesis | Vesicoureteric reflux grade V | Urethral stenosis |
| Unilateral absent kidney | Bilateral ureteropelvic junction obstruction | Urethral stricture |
| Unilateral polycystic kidney | Bilateral ureterovesical junction obstruction | Agenesis of distal urinary system: bladder/urethra |
| Unilateral dysplastic kidney | Bladder exstrophy | Bilateral dysplastic kidneys |
| Unilateral multicystic kidney | | Autosomal recessive polycystic kidney disease (ARPKD) |
| Horseshoe kidney | | Bilateral renal agenesis |
| Pelvic kidney | | |
| Duplicated kidney | | |
| Duplicated renal pelvis | | |
| Duplicated ureter | | |
| Vesicoureteric reflux grade I | | |
| Vesicoureteric reflux grade II | | |
| Unilateral ureteropelvic junction obstruction | | |
| Unilateral ureterovesical junction obstruction | | |
| Autosomal dominant polycystic kidney disease (ADPKD) | | |

**Supplemental File 7:** Results of Real-Time Clinical Model Simulation

| Operational Point (Avg. % of Patients with AKI alert) | Avg. # of Patients with AKI Alert | % of AKI Predicted | Positive Predictive Value [%] | PPV Lift |
|---|---|---|---|---|
| 5% | 3.6 | 38.0% | 3.6% | 5.4 |
| 15% | 10.6 | 57.8% | 2.2% | 3.2 |

*Results of a real-time clinical simulation of the ideal LASSO model for to potential intervention-levels.*
